# Factor Structure, Reliability and Validity of the Institutional Culture Assessment Scale (ICAS)

**DOI:** 10.1101/2020.09.28.20203331

**Authors:** Gabriela Massaro Carneiro Monteiro, Carolina Meira Moser, Luciana Terra de Oliveira, Glen Owens Gabbard, Pricilla Braga Laskoski, Simone Hauck

**Author notes:** **Corresponding author:** Gabriela Massaro Carneiro Monteiro, Pós-graduação em Psiquiatria e Ciências do Comportamento, Faculdade de Medicina, Universidade Federal do Rio Grande do Sul, Rua Ramiro Barcelos, 2400, Porto Alegre, RS, Brazil, CEP: 90035-903. **Data Availability Statement:** The data that support the findings of this study are available from the corresponding author upon reasonable request.

## Abstract

**Introduction:** Work environment can affect the employees, fostering well-being versus emotional burden. The aim of this study was to develop the Institutional Culture Assessment Scale (ICAS), and evaluate its Factor Structure, Reliability and Validity in a Brazilian sample of medical students and physicians in different settings and phases of the medical career.

**Method:** 2,537 individuals were evaluated by an online questionnaire. The sample was split in half for independent testing of Exploratory Factor Analysis and Confirmatory Factor Analysis. We then used Confirmatory Factor Analysis (CFA) to test the best solutions in the second half of the sample. Then, considering a unidimensional model solution, an item response theory (IRT) analysis was conducted. Simple linear regression analysis was performed to investigate associations between ICAS factor scores and internal validators (burnout scores), using again the second half of the sample.

**Result:** Parallel analysis revealed two factors. The first factor encompassed items involving the institution and supervisors. The second factor encompassed items involving peers. We decided to performed the next analysis with a unidimensional construct based solely on institution/supervisor items. A unidimensional model including the remaining seven items from the ICAS instrument revealed an excellent fit with the data. All items loaded significantly on the unidimensional latent trait with factor loadings ranging from 0.583 to 0.869. McDonald’s Omega was 0.89, showing a high internal consistency.

**Conclusion:** This study presents a valid and reliable scale to assess aspects of institutional culture connected to the relationships with superiors/supervisors and to the relation to the institutions themselves.

## Introduction

It is now a well-established axiom within the organizational literature that the institutional environment plays a key role in the efficiency, health and well-being of professionals. Institutional culture subtly shapes attitudes, biases, and behaviors, defining what is encouraged, discouraged, accepted or rejected within the workplace or profession. However, the essence of institutional culture, sometimes referred to as organizational culture, is a somewhat elusive and complex construct that defies simple definition. There are a myriad of formal definitions and a variety of models and methods for assessing it, some of which are likely to be found in the human resources field, where the focus is mainly on performance evaluation. Most of the scales assess the degree of satisfaction with the perceived climate at work in terms of self-realization, workload, co-workers, salary structure, leadership, nature of the work, promotions, and other markers in an effort to catalog the multiple factors that make up the structure. The end result is a more global assessment of the work environment, one that is less focused on factors related to employees’ well-being and psychological health.^1–4^

The institutional culture may be of particular importance to those who are health care professionals. Several authors have observed that health professionals as a group have been intensely affected by changes in the administrative structure of hospitals and the decline of physician autonomy in the current culture of health institutions.^5–7^ In fact, a recent meta-analysis revealed a significantly smaller effect of interventions aimed at the individual level compared to those geared to the institutional level.^8,9^ Rates of exhaustion and mental health concerns are currently affecting 30-60% of all doctors and physicians in training in North America.^10^ In the face of the COVID-19 pandemic, these rates can be expected to rise exponentially.^11–13^

Adequate tools to identify the specific aspects in the culture that contribute to staff well-being versus those that result in distress and psychiatric symptoms are badly needed. Although some instruments exist that measure the link between good relationships and work-related variables^14^, to our knowledge, there is not an instrument that evaluates the relationships with others that exist within the institution as well as the leadership style along with the relation of the professionals to the institution itself (e.g., the feeling of belonging, a collaborative atmosphere and an alignment between the values of doctors/ health professionals and the institution). Those factors seem to be highly relevant for the configuration of the culture, in that they may be instrumental in preventing an erosion of the professionals’ role, their psychological health, the quality of service provided to the clients, and in the case of health professionals the quality of patient care, besides the cost to society.^15^

With the intention of identifying these aspects Carneiro Monteiro et al.^16^ developed the Work Environment Evaluation Instrument (WEEI), to evaluate the work environment in medical training settings. The WEEI was developed and tested in a specific population, i.e., residents in training. The aim of the present study was to develop the Institutional Culture Assessment Scale (ICAS), based on an adaptation of the WEEI’s items, testing its factor structure, reliability and validity in a Brazilian sample of medical students and physicians in different settings and phases of the medical career.

## Methods

In this online cross-sectional study, 2,537 individuals were evaluated in October 2019, using a snowball method over fifteen days. We chose an electronic questionnaire in order to reach a greater number of respondents. Online questionnaires also have the potential advantage of enhancing reliability by augmenting the perception of anonymity.^17,18^ The advertisement explicitly stated the survey’s anonymity, and the subject only continued to the questionnaire if agreed with the online informed consent form. After completion, the questionnaire provided telephone and electronic contact information for suicide prevention and support centers located in Brazil. The study was approved by the ethics committee of the Hospital de Clínicas de Porto Alegre (Porto Alegre, Brazil) (protocol 70231617.6.0000.5327).

### Sample

All 26 Brazilian States and the Federal District were represented in the sample, although some Brazilian regions were more heavily represented than others: 48% of participants were from the Southern region, 28.1% from the Southeast region, 5.6% from the Midwest region, 15.2% from the Northeast region and 2.4% from the Northern region. This difference probably reflects the advertising strategies, and to some extent the distribution of doctors in Brazil. The mean age was 39.92 (SD 12.64). Sociodemographic and work-related variables are described in table 1.

**TABLE 1.**
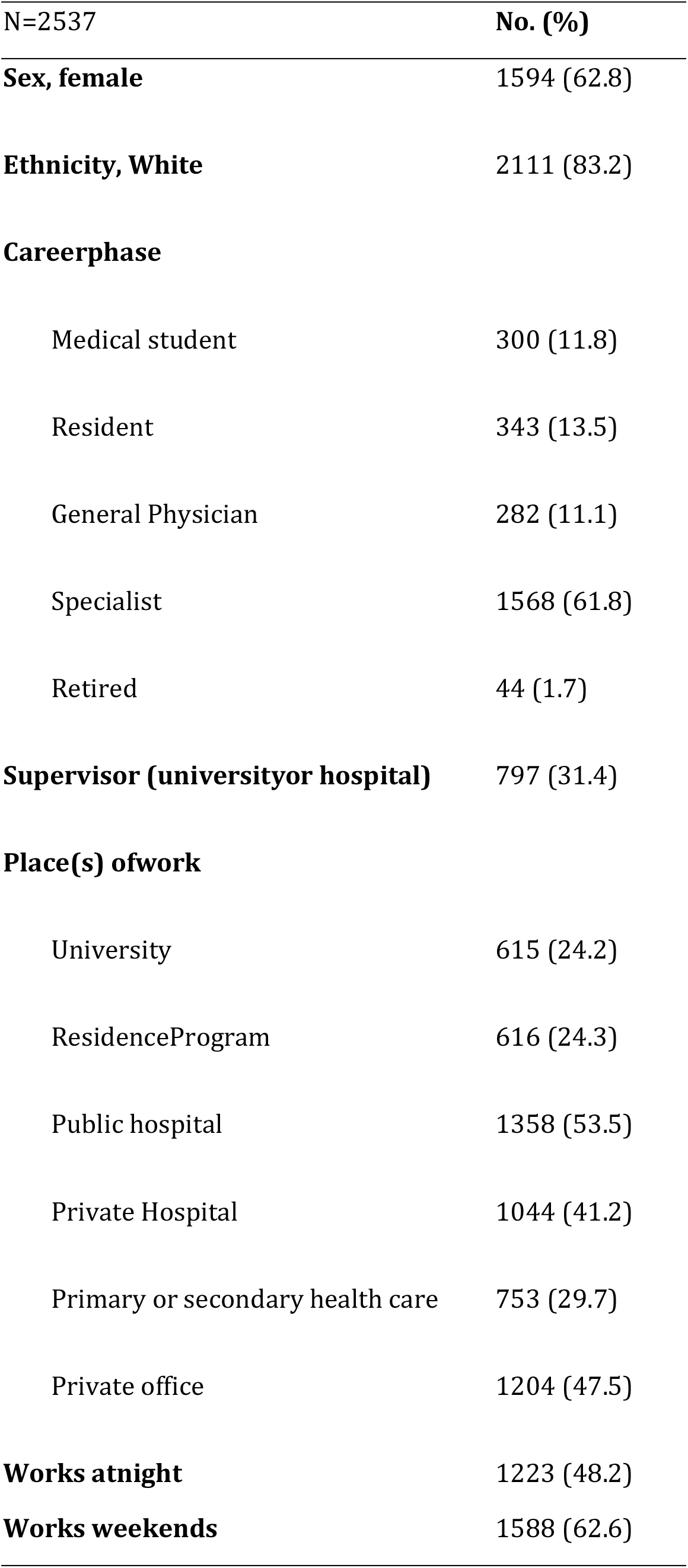
Description of the Sample

### Survey Instruments

Besides sociodemographic and work-related information, the online questionnaire included the following instruments used in this study:

#### The Work Environment Evaluation Instrument (WEEI)^16^ – adapted version

The WEEI was developed based on several focus groups with experts, professors, medical students and residents. The aim was to elaborate on the items related to the work environment that could be related to emotional distress in training physicians’ populations. At first, fourteen items were elaborated and tested in a pilot study.^19^ The comprehension of the items and the face validity were evaluated and discussed, and small adjustments were made. The resultant version, called WEEI, has 11 items. It is a Likert scale of five points where 0 corresponds to “Totally false” and 4 corresponds to “Totally true”. Five items evaluate the relationship with preceptors/supervisors, three with colleagues/peers and three the relation to the institution itself. The instrument evaluate aspects such as feeling comfortable asking for help, feeling heard and helped versus feeling pressured by preceptors/supervisors, the feeling of belonging and the presence of a collaborative atmosphere in the institution, the perception of support by peers, among others. The WEEI presented high internal consistency and reliability: the Cronbach’s Alpha coefficient (α) for the total score in the validation study was 0.898, and for each of the dimension it was 0.79 for “institutions”, 0.78 for “colleagues”, and 0.87 for “preceptors”. Both total scores and dimensions correlated significantly with Burnout scores (p<0.01).^16^ For the current study the items were adapted to be suitable for all phases of medical career and tested in a national sample of physicians and medical students with the aim of developing the ICAS.

#### Maslach Burnout Inventory – Human Services Survey (MBI-HSS)

The MBI-HSS measures burnout on three subscales: emotional exhaustion (EE), depersonalization (DP), and the sense of personal accomplishment (PA). It is a self-completion questionnaire answered by a Likert scale of seven points with “0” being“never” and “6” every-day. These three dimensions are related to each other but independent.^20^

### Statistical Analysis

First, the sample was split in half for independent testing of Exploratory Factor Analysis and Confirmatory Factor Analysis. Second, all items from the WEEI adapted version entered an Exploratory Factor Analysis (EFA). Parallel analysis compares the scree of factors of the observed data with that of a random data matrix of the same size as the original providing the best number of factors in the sample using polychoric correlations to account for the categorical nature of the items. EFA was performed using the weighted least square factoring method for polychoric correlations.

Third, we used Confirmatory Factor Analysis (CFA) to test the best solutions in the second half of the sample. For fit indices, the following indices were estimated: comparative fit index (CIF), Tucker-Lewis index (TLI), and root mean square error of approximation with 90% confidence interval (RMSEA-90%CI). A CFI and TLI values about 0.90 or close to 0.95 represent a good fit. RMSEA values close to or below 0.05 represent a good fit, and those below 0.08 an acceptable fit. Moreover, the reliability of the best factor solutions was tested by means of the McDonald’s Omega.

Fourth, considering a unidimensional model solution, an item response theory (IRT) analysis was conducted using the graded response model in the second half of the sample.^21^ The latent trait that we called “institutional culture” was centered on 0 with a SD of 1. The importance of using also IRT to analyze the scale has several reasons. One of them is that we want to know if the instrument is evaluating well both sides of the latent trait. For instance, we need to know if it captures well both healthy and toxic institutional environments. That is important because, besides understanding what contributes to developing mental problems, it is also very important to know what promotes health and well-being within the work environment. Also, it is important to evaluate the performance of each item in separate and how they individually contribute to the measurement of the latent trait. This is essential, not only, but also to evaluate if a simple sum of the items would be a sufficiently reliable measure of the latent trait, namely “institutional culture”, to be used in further studies.

Finally, simple linear regression analysis was performed to investigate associations between ICAS’ factor scores and internal validators (burnout scores) using again the second half of the sample.

Parallel analysis and Exploratory Factor Analysis were performed using the *fa*.*parallel()* and fa functions from the psych package. ^22^ Confirmatory Factor Analysis was performed using the *cfa*() function from the lavaan package.^23^ Item response Theory Analysis was performed using the *grm()* function the ltm package^24^ and *irt*.*fa()* function from the psych package.^22^ Regression analysis were performed with the *lm()* function from base R. All analyses were performed in R.^25^

## Results

### Parallel analysis and Exploratory Factor Analysis

Parallel analysis revealed that 2 factors represents the best solution to explain variance in the WEEI adapted version items (Figure 1). The first factor explained 36% of the variance, whereas the second explained 15% of the variance. The first factor encompassed items involving the institution (Table 2; items 2, 4, 6) and supervisors (Table 2, items 1, 3, 7, 9). The second factor encompassed items involving peers (Table 2, items 5, 8, 10). The item 11 presented a factor loading which is markedly lower than the average of the factor loading from the other items of the first dimension (λ=0.23 vs. average λ=0.73). The correlation between the first and second factors was moderate 0.47.

**Table 2.** Exploratory Factor Analysis Results from the first half of the sample (n=1269)

| Item code | Item content | Factor 1 | Factor 2 | Communalities | Uniquenesses |
| --- | --- | --- | --- | --- | --- |
| 1 | I feel more pressured than helped by my superiors / supervisors | **0,7** | -0,07 | 0,44 | 0,56 |
| 2 | I feel like I belong to my institution. | **0,6** | 0,05 | 0,39 | 0,61 |
| 3 | I am not afraid to ask for help from my superiors / supervisors. | **0,67** | -0,08 | 0,41 | 0,59 |
| 4 | I feel a collaborative climate in my institution. | **0,83** | 0,01 | 0,7 | 0,3 |
| 5 | I talk to my colleagues about problems at work / college, and this helps me deal with everyday problems. | 0,22 | **0,46** | 0,35 | 0,65 |
| 6 | I think the values of my institution are in accordance with my own values. | **0,72** | 0,04 | 0,55 | 0,45 |
| 7 | I feel helped by my superiors / supervisors. | **0,85** | 0,02 | 0,74 | 0,26 |
| 8 | I feel that colleagues are just colleagues and do not get involved with my problems. | -0,01 | **0,83** | 0,68 | 0,32 |
| 9 | My superiors / supervisors understand and listen when I have complaints. | **0,75** | 0,02 | 0,58 | 0,42 |
| 10 | My colleagues are not my friends. | -0,02 | **0,81** | 0,64 | 0,36 |
| 11 | I feel that I am always short of what the superiors / supervisors expect of me. | 0,35 | -0,01 | 0,12 | 0,88 |

Note: Factor loadings higher than 0.4 are marked in bold; the items were translated to English from the original Brazilian Portuguese Version

**Figure 1.**
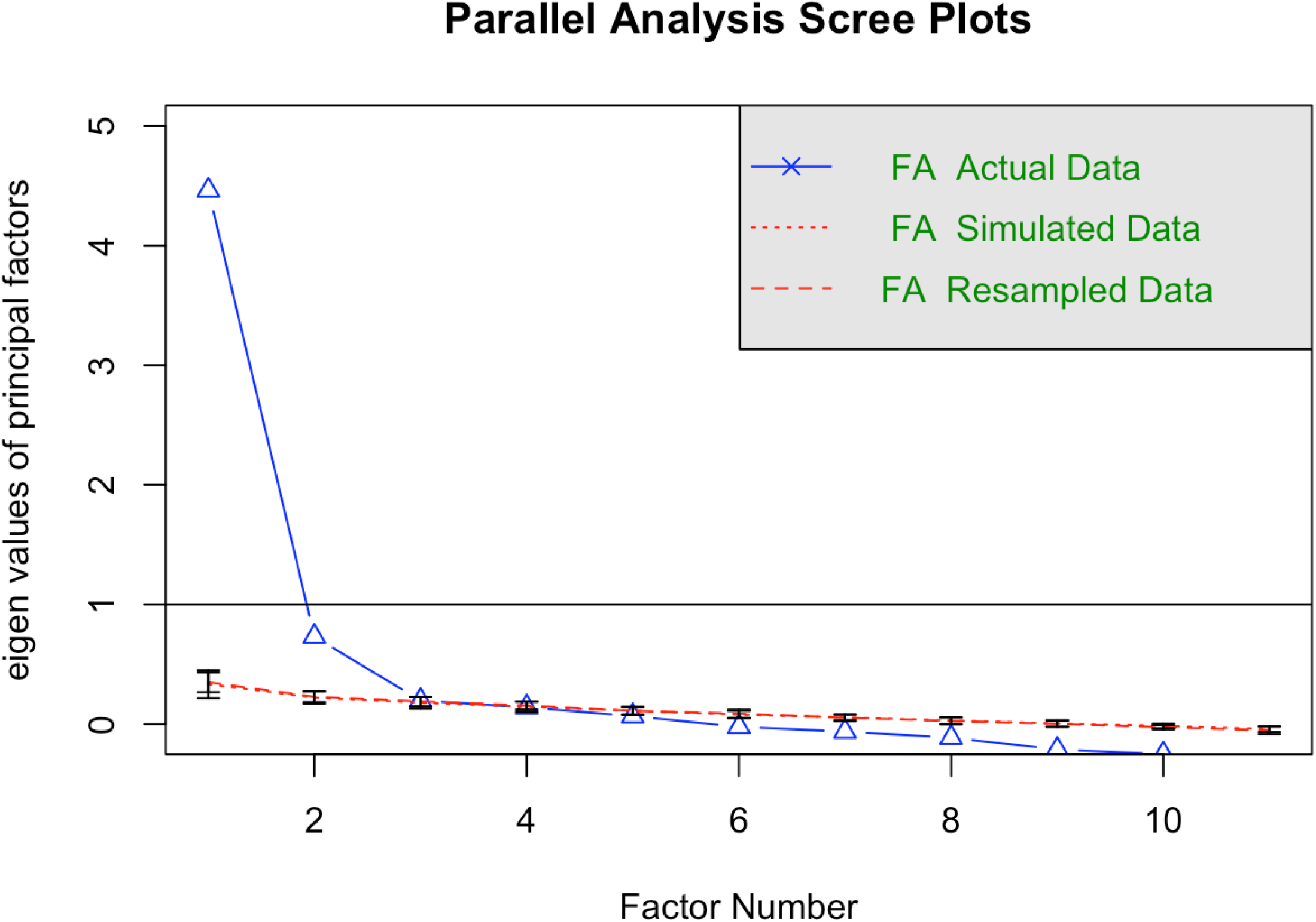
Parallel Analysis Scree Plots for determining the number of factors in Exploratory Factor Analysis.

Because peers formed a second factor, separated from institution and superiors/supervisors’ factors and presented with only 3 items, we decided to exclude the peer domain and performed the next analysis with a unidimensional constructed based solely on institution/supervisor items.

### Confirmatory Factor Analysis and Internal Consistency

A unidimensional model including the remaining 7 items from the ICAS revealed an excellent fit to the data (RMSEA=0.053, CFI=0.997, TLI=0.996). All items loaded significantly on the unidimensional latent trait with factor loadings ranging from 0.583 to 0.869. McDonald’s Omega was 0.89, showing a high internal consistency. (Table 3)

**Table 3.** Confirmatory Factor Analysis and Item Response Theory Parameters

| CFA |  |  |  | IRT |  |  |  |  |  |  |  |  |
| --- | --- | --- | --- | --- | --- | --- | --- | --- | --- | --- | --- | --- |
| Items | Factor Loadings |  |  | Thresholds |  |  |  | Disc | Difficulties |  |  |  |
|  | Estimate | SE | p-value | 1 | 2 | 3 | 4 | a | b1 | b2 | b3 | b4 |
| WEEI2 | 0,583 | 0,021 | <0.001 | -1,398 | -0,933 | -0,421 | 0,684 | 1,471 | -2,077 | -1,265 | -0,574 | 1,045 |
| WEEI4 | 0,816 | 0,012 | <0.001 | -1,152 | -0,528 | -0,131 | 1,006 | 2,878 | -1,305 | -0,587 | -0,169 | 1,164 |
| WEEI6 | 0,767 | 0,014 | <0.001 | -1,05 | -0,456 | 0,049 | 1,114 | 1,983 | -1,386 | -0,578 | 0,068 | 1,549 |
| WEEI1 | 0,659 | 0,019 | <0.001 | -0,759 | 0,123 | 0,57 | 0,954 | 1,515 | -1,074 | 0,127 | 0,801 | 1,479 |
| WEEI3 | 0,631 | 0,02 | <0.001 | -1,318 | -0,727 | -0,38 | 0,376 | 1,402 | -2,137 | -1,16 | -0,654 | 0,57 |
| WEEI7 | 0,869 | 0,01 | <0.001 | -1,152 | -0,558 | -0,093 | 1,05 | 3,051 | -1,293 | -0,626 | -0,087 | 1,198 |
| WEEI9 | 0,766 | 0,014 | <0.001 | -1,039 | -0,501 | -0,013 | 1,171 | 2,103 | -1,376 | -0,55 | 0,052 | 1,584 |

Note: CFA - Confirmatory Factor Analysis; IRT - Item Response Theory; WEEI - Work Environment Evaluation Instrument.

### Item Response Theory

Graded Response Model revealed high discrimination for all items (all above 1.4) and difficulties ranging from −2.1 for the lower threshold to 1.5 for the higher threshold. Test information function (Figure 2, Panel A) reveals the test captures reliable information from −2 (alpha 0.8) until +2 area (alpha 0.72) of the latent trait as revealed by the irt.fa function and has a good potential in differentiate both healthy and toxic environments. Item Information Curves (Supplemental Figure 1) and Item Characteristic Curves are (Supplemental Figure 2) are presented in supplemental material. The ICAS showed approximately normal distributions in summed and IRT-based scores (Figure 2, Panel B). Conversion tables between summed and IRT-based scores are presented in Supplemental Table 1.

**Figure 2.**
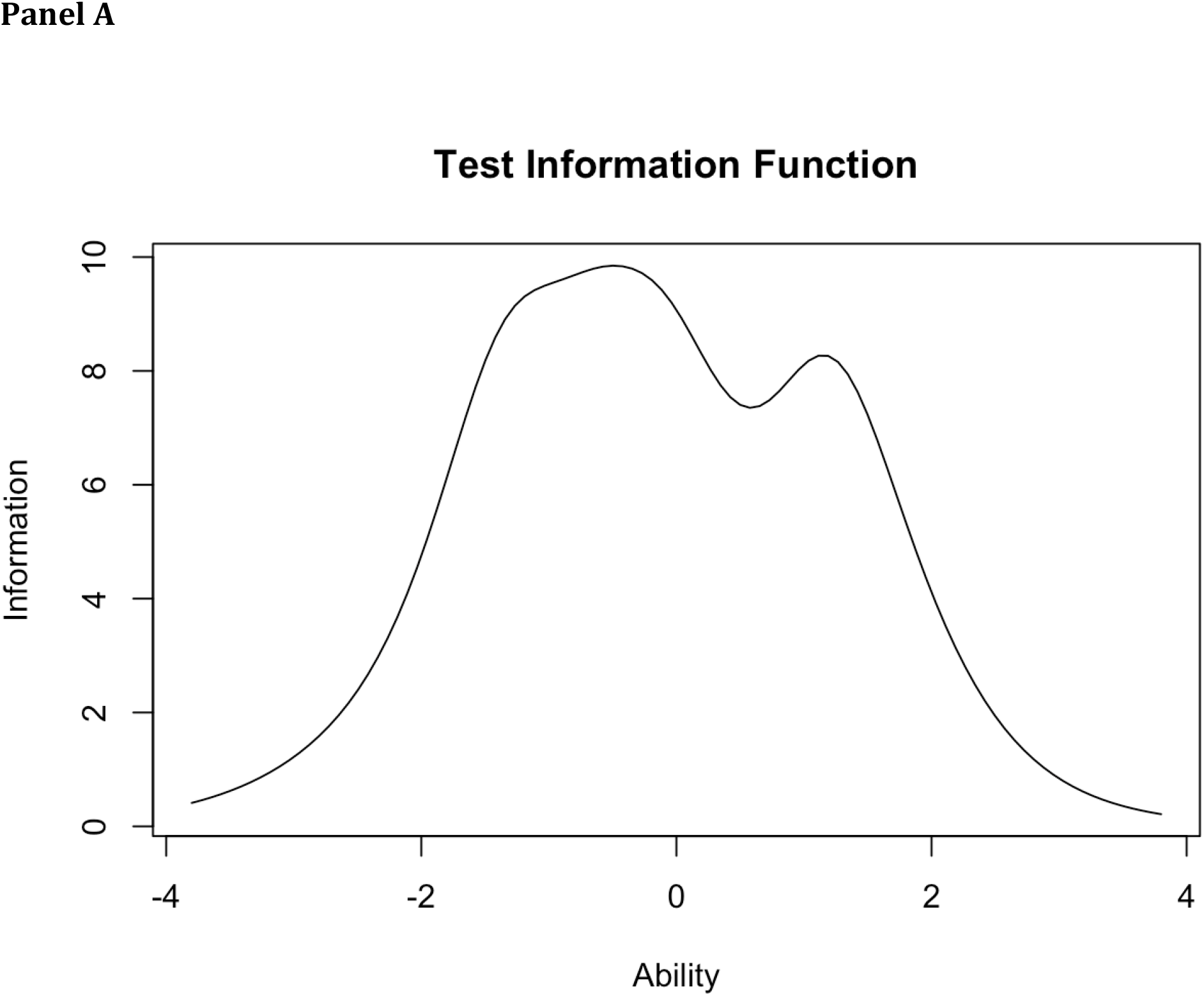
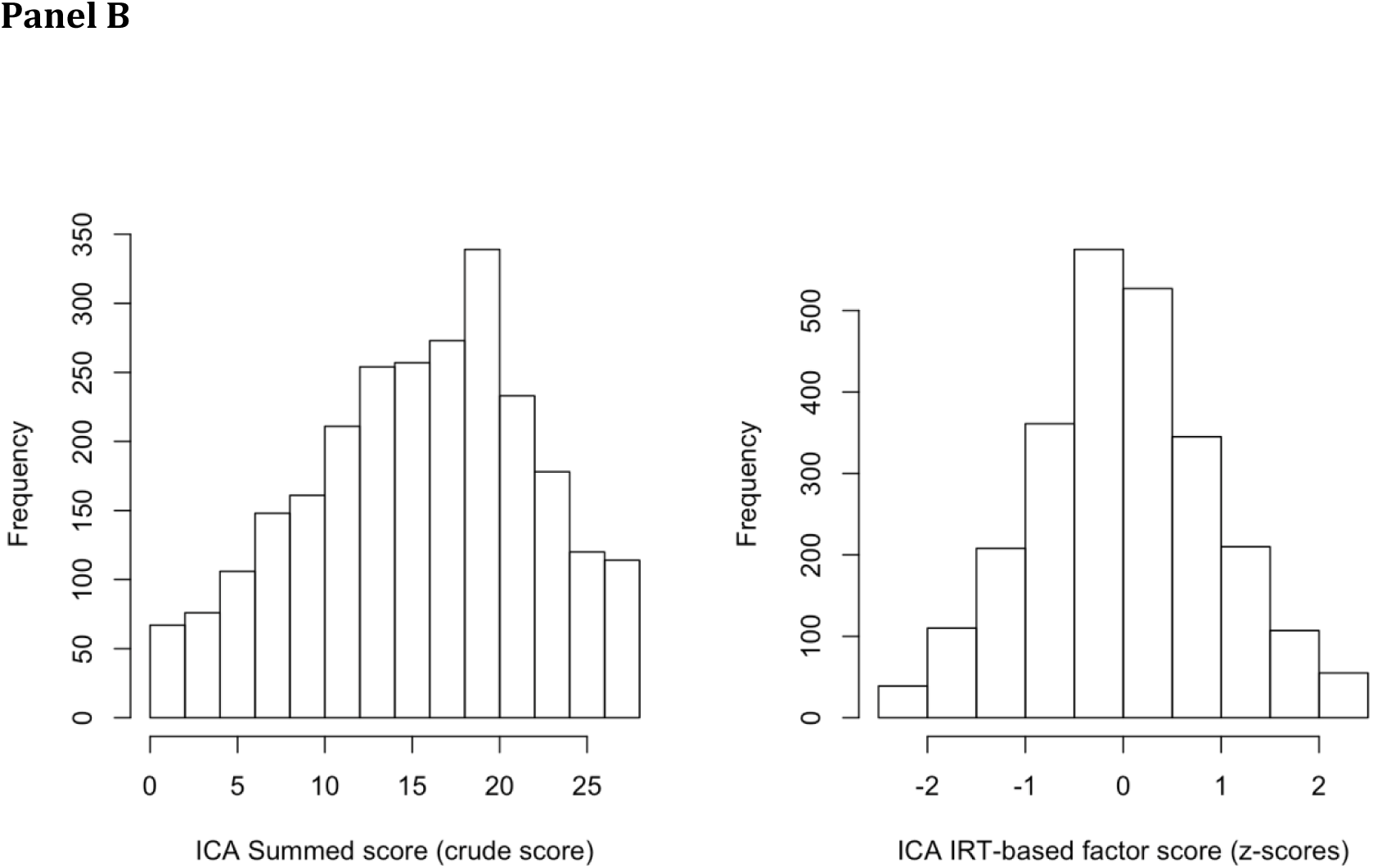
Test Information Function for the revised structure of the ICA and histograms of the summed and IRT-based factor scores.

### Construct and external validty

Finally, ICAS factor scores explain 22% of the variance of the Emotional Exhaustion score (p<0.001), 10% of the Depersonalization score (p<0.001) and 14% of the Personal Achievement score.

## Discussion

This study presents a valid and reliable scale to assess institutional culture aspects connected with the relationships with superiors/supervisors and the relation to the institutions themselves. The ICAS revealed an excellent fit of the data (RMSEA=0.053, CFI=0.997, TLI=0.996) as well as high internal consistency (McDonald’s Omega = 0.89), proving to be a suitable tool for the evaluation of institutional culture dimensions that might be related to staff well-being versus those that result in distress and psychiatric symptoms. The correlation with burnout score in this sample provide additional evidence in this sense. The ICAS also encompasses many concepts of self-determinant theory research that focuses on the consequences of the extent to which individuals are able to satisfy the needs within social environments.^2^

To the best of our knowledge, there isn’t an instrument that evaluates the relationships with superiors/supervisors along with the relation to the institution itself as dimensions of the institutional culture that may impact employees’ health. There are some instruments like the Working Environment Scale (WES-10)^26^ that assesses an overall degree of satisfaction with the perceived climate at work in terms of self-realization, workload, conflict and nervousness. Another is the Organizational Culture Assessment Instrument (OCAI)^27^ that aims to evaluate institutional culture regarding six attributes: dominant characteristics, organizational leadership, management of employee, organization glue, strategic emphases, and criteria of success. The ICAS has the advantage of being a brief self-applied instrument that takes into account the perception of individuals in different stages of career and the holding of different positions within the institution. The items evaluated have been proven to be related to burnout symptoms in previous studies with medical residents as well in this national sample of medical studies and physicians.^16,19^ As the aspects evaluated are related to leadership style (e.g. fostering collaboration, emphatic listening and attitudes towards employees), and to the perception of the institution (e.g., collaborative climate, shared values), the ICAS is expected to be associated with employee mental health in populations other than health professionals and should be tested in different settings. It is important to point out that the aspects evaluated by the ICAS are possible to change by interventions in the institutional level. This approach can be a more effective strategy than limiting the scope to interventions at the individual level that tend to foster the augmentation of the individual resilience, but, in many cases, ignore some of the causes.^7,8,28^

It is well established that the work environment plays an important role in subjects’ well-being and/or distress. However, the aspects of the institutional culture that are more related to psychological distress in the work setting are still not defined. A variety of factors, e.g., individual vulnerability, institutional pressures, and socioeconomic factors, are involved in the phenomena that lead individuals to become severely distressed in the context of the workplace.^5,29,30^ When individual factors interact with institutional characteristics that reinforce the unhealthy aspects of the individual (such as an excess of competitiveness, criticism, low self-esteem and so forth), there can be devastating health consequences.

This investigation has notable strengths. First, we were able to develop a fast and self-administered instrument with high internal consistency that evaluated institutional environment in a sample of medical students and physicians. Second, we were able to test the instrument in a high number of participants. Third, we could evaluate aspects that are poorly addressed in the literature concerning the work environment features. Finally, we found an association between the ICAS score and burnout dimensions, reinforcing its validity in measuring aspects related to work related distress vs. well-being. On the other hand, this study has limitations typical of preliminary studies that need replication in other samples and contexts. On the other hand, this study has limitations inherent to preliminary studies that need replication in other samples and contexts. Also, it used a convenience sample in an online study, so only participants with easy access to internet were likely and able to participate in the present study.

Several authors have emphasized that changing the culture within institutions may be the most efficient way to stem the rising rates of emotional distress among health professionals.^7^ In the case of health professionals, a recent paper by Hartzband and Groopman^6^ call the attention to the fact that the modifications implemented in the health systems aiming at rising productivity and efficiency have eroded the very sense of the identity of being a doctor by taking away the important triad of autonomy, competence and relatedness, described by Gagne and Deci^2^ as essential to support the professional’s intrinsic motivation and psychological well-being. This might well be the case in other professions where the sense of effectiveness has followed an excessive materialistic view at the expense of essential human emotional needs. In this sense a validated and reliable instrument like the ICAS may help in calling the attention to those aspects that seem to be essential to foster well-being within any institution.

## Data Availability

The data that support the findings of this study are available from the corresponding author upon reasonable request.

## Acknowledgments

We thank Professor Giovanni Abrahão Salum Júnior for the assistance in the statistical analyses and we thank Professor Fernanda Lucia Capitanio Baeza for the assistance in the development of the Work Environment Evaluation Instrument (WEEI).

